# Reducing the use of sleep-inducing drugs during hospitalization by a multi-faceted intervention – a pilot study

**DOI:** 10.1101/2021.10.06.21264630

**Authors:** Stephanie Heinemann, Jonas Klemperer, Eva Hummers, Roland Nau, Wolfgang Himmel

## Abstract

**Objectives:** Many patients receive benzodiazepines or Z-drugs during hospitalization due to sleeping problems. In a pilot study, we aimed to find out whether, and to what degree, a multi-faceted intervention can reduce the use of these drugs, especially in older patients and those without a psychiatric or neurological disorder. The results of this pilot study should inform the design of a randomised controlled trial (RCT).

**Methods:** In a quasi-experimental design, we implemented the intervention in a German hospital with the support of the hospital director, medical and nursing staff and employee representatives. We compared prescription data for sleep-inducing drugs before and after the intervention by Fisher’s exact test and used Odds-Ratios (ORs) with their 95 % confidence intervals (CI) as measure of effect size.

**Results:** The data from 960 patients aged 65 and older before intervention and 1049 patients after intervention were analysed. Before intervention, 483 (50.3%) of the patients received sleep-inducing drugs at some time during their hospital stay. After the intervention, 381 (36.3%) patients received a sleep-inducing drug, resulting in an OR of 0.56 (95% CI: 0.47 to 0.68; p <0.001). The reduction was particularly pronounced in patients without a psychiatric or neurological disorder (from 45.0% to 28.8%). Especially benzodiazepines were significantly reduced (from 24.3% to 8.5%; OR: 0.31 (0.23-0.4); <0.001).

**Conclusions:** A multi-faceted intervention to change the practice of the use of sleep-inducing drugs in one hospital was successful in terms of drug reduction, particularly for benzodiazepines. The intervention was effective especially for target persons, i.e. those without a psychiatric or neurological disease. Being aware of the magnitude of the change and the role of important stakeholders could help researchers, hospital physicians and hospital pharmacists to design a large RCT, including control hospitals, to evaluate the success of a multi-faceted intervention on a scientifically sound basis.

**KEY MESSAGES:** *What is already known on this subject:*

- Benzodiazepines and Z-drugs are still too often used for sleep problems in hospitals.
- Simple interventions, such as training seminars to reduce the use of benzodiazepines, Z-drugs, and other drugs for insomnia treatment are limited in their effect.

*What this study adds:*

- The intervention significantly reduced the rate of sleep-inducing drugs by 14 percentage points.
- The results of this pilot study give a first impression of the possible impact of the interventions and provide essential information for the design of a randomized controlled study.

## INTRODUCTION

Many patients have trouble sleeping in the hospital due to environmental factors such as unfamiliar sounds, nursing interruptions, uncomfortable beds and bright lights and probably also due to worries and fears [1]. Although these sleep problems do not fulfil the clinical diagnosis of insomnia [2], they are often treated with benzodiazepines and newer non-benzodiazepines, so-called Z-drugs [3]. While these drugs may help patients to sleep in the hospital environment, they also have adverse effects, such as confusion, falls, fractures and craving, especially in older patients [4].

A recently published review showed that simple interventions (such as training seminars) to reduce the use of benzodiazepines, Z-drugs, and other drugs for insomnia treatment are limited in their effect [5]. Some of the multi-faceted interventions studied in this review were efficient, especially when healthcare professionals and patients were actively involved. The majority of these studies was based on small patient samples, selected hospital wards and short intervention periods. Consequently, we considered it important to expand our knowledge of what works by a larger intervention.

Together with all relevant stakeholders, we developed a tailored intervention, called “The Sleep-friendly Hospital Initiative” [7], with the ultimate goal of reducing the use of sleep-inducing drugs in hospitals or making their use more appropriate (for more details of this initiative, see the Methods section). As a first step, we needed to better understand this drug use in the hospital setting— for two reasons:

1. The use of drugs in hospitals follows only partly pharmacological criteria; their use is also a matter of non-medical or context factors, as Helman states in his early research [19], or as a matter of games with specific rules and strategies within the habitus of the social world of a hospital, as Bourdieu [20] put it.
2. Any attempts to interrupt this smooth-running game begin by showing all relevant stake-holders that we understand what is going on in the hospital and what the reasons for their performance are. Only then will it be possible to develop together with them new and reliable rules of how to cope with transient sleep problems in the hospital.

The project followed the Medical Research Council (MRC) framework for designing and evaluating complex interventions to improve health care [21, 22]. One aim of this framework is to ensure that interventions are empirically and theoretically founded. Therefore, we explored the real extent of the use of sleep-inducing drugs in the hospital where the intervention should take place and the reasons for their use as well as the experience with these drugs, seen from the perspective of doctors, nurses and patients. We chose a mixed-methods approach to collect the data needed. As a first step, we studied the use of sleep-inducing drugs for transient sleep problems in one hospital[6], including a chart review of the patient hospital files, a survey of doctors’ and nurses’ use of, and experience with, sleep-inducing drugs and a standardized patient survey. The goal of these studies, both individually and collectively, was to understand the current practice and to identify possible changes that could improve this practice.

Reviews showed that formal didactic conferences and passive forms of medical education, such as brochures or printed clinical guidelines are the least effective methods for changing physician behaviour [23]. Moreover, stakeholder engagement is essential for moving knowledge into action within healthcare [24]. This is the heart of the MRC framework. Based on our results, we identified several areas with a potential for improvement and worked together with the stakeholders of the hospital to create an intervention strategy and to implement a multi-faceted hospital intervention. The different facets of the intervention correspond to what Atkins et al. have identified as domains which influence behaviour change, such as knowledge, skills, social/professional role, environmental context and resources [7].

The aim of this pilot study was to find out whether our multi-faceted Sleep-friendly Hospital Initiative (1) reduced the use of sleep-inducing drugs, such as benzodiazepines and Z-drugs, (2) was especially effective in patients without a psychiatric or neurological disorder and avoided an increased prescription of sedative anti-depressants or anti-psychotics (which may be considered by hospital personnel as an alternative strategy). The results of this pilot study should inform a larger randomized controlled trial.

## METHOD

### Study design

In a quasi-experimental design, we implemented a multi-faceted, tailored intervention [8] to reduce the use of sleep-inducing drugs for older patients in a German hospital over a period of 18 months (October 2016 to June 2017). To measure the success of the intervention, we compared prescription data before the begin of the study period (July-August 2013) and after the intervention period (July-August 2017).

### The Sleep-friendly Hospital Initiative

The initiative incorporates a strategy of making appropriate decisions about sleep-inducing drugs during the day in order to replace ad-hoc prescriptions during the night. The strategy consists of three main elements: (1) For patients who sleep well at home, the initiative encourages doctors and nurses to treat hospital-associated sleeping problems with non-drug options. (2) Only if these options fail, should sleep-inducing drugs be ordered as a p.r.n. prescription (from Latin: *“pro re nata”*; meaning “as needed” or “as the situation arises”), especially valerian, mirtazapine, melperone or zolpidem in low doses as recommended by the German PRISCUS list [9]. (3) For patients who regularly use sleep-inducing drugs before hospital admission, the initiative encourages hospital physicians to critically assess this prescription, making changes carefully to avoid withdrawal symptoms.

### Hospital

The intervention hospital is a mid-sized regional German hospital (485 beds) with departments of internal medicine, geriatrics and several surgical sub-specialties.

### Data collection

Anonymous data from the hospital charts of all patients 65 years and older who spent at least one night in the hospital (and not in intensive care) was extracted for analysis. Data was collected from two time points, separated by exactly four years. The first time point was before the project began, i.e. before any studies described in the study protocol [6] were carried out in the intervention hospital. The second time point followed the 18-month-long intervention phase which included several components described in detail in Chapter 5 of Heinemann 2020 [8].

Patients that died during their hospital stay were excluded. We assumed that an increased amount of care was necessary in these cases that could go hand in hand with an increased use of sleep-inducing drugs. Another exclusion criterion was a stay that was spent entirely on the intensive care unit. Drugs that were administered on an Intensive Care Unit were not counted in the data collection.

### Drugs

Drug data included both the pharmacological class of drug and how often it was administered (number of days). We defined which drugs are sleep-inducing drugs, inspired by several authors [10–13]. Our list included anti-depressants, anti-psychotics, benzodiazepines, Z-drugs and valerian (**Error! Reference source not found**.).

### Data extraction

Information on patients and their medication was not electronically available in the hospitals. We manually extracted the data from the patient charts into a databank. Patient charts varied in their order and length. Four advanced medical students and one nurse extracted the data and entered it into the study databank. To ensure data quality, we developed a standard operating procedure for data extraction and data entry.

Demographic data included sex, age and where the patient was admitted to the hospital from (home, nursing home, other hospital, other department within the same hospital).

Hospital data included length of hospital stay and department (internal medicine, surgery, geriatrics) where the patient was treated.

Medical data included whether or not the patient had specific psychiatric/neurological diagnoses (such as dementia, depression, panic, insomnia, etc.) and if the patient reported a current prescription for sleep-inducing drugs at the time of admission.

Drug data included both the type of drug and how often it was administered (number of days).

### Outcomes

Primary outcome was the percentage of patients receiving any sleep-inducing drug at any time during their hospital stay. The use of specific classes of sleep-inducing drugs were analysed as secondary outcomes.

### Sample-size calculation and statistical analysis

In a combined patient survey and chart review from the same hospital[14], 21% of the older patients had at least received one benzodiazepine during their hospital stay. We intended to lower this rate by 5 percentage points. To detect a significant reduction with an alpha of 5% and a power of 80%, we needed a sample of N = 982 patients in both time periods [15].

Fisher’s exact test was used to investigate whether the change between these periods was significant. Odds-Ratios (ORs) with their 95 % confidence intervals (CI) were used as measures of effect size. All analyses were performed with SAS, version 9.4.

### Re-analysis of a sub-sample of data

To assess how accurately we conducted our first data collection and how consistently we trained the data extractors, we drew a subsample of 220 patients and then compared the entries of the two data sets. The patient files for this confirmatory quality analysis were extracted and assessed by a person that had not been involved in the first data collection in this hospital and had no prior contact to that hospital and its patient files. Agreement between the first and second entry was assessed by the kappa-statistic [16].

There was no difference in the classification of the patient’s gender between the first and second entry. Only in 3 cases (1.4%), the two entries differed in the classification of whether a patient had received a sleep-inducing drug or not, yielding a kappa of 0.97. In 7 cases (3.2%), the entries differed on whether a patient received a potentially inappropriate medication or not (kappa: 0.88). In 26 cases (11.8%), the patient’s diagnoses were entered differently (kappa: 0.71). Kappa values above 0.61 are interpreted as substantial agreement, above 0.81 as almost perfect agreement [16].

## RESULTS

### Implementation of the Sleep-friendly Hospital Initiative

The different recommendations of the Sleep-friendly Hospital Initiative were communicated over several avenues to doctors, nurses and patients. They were visible hospital-wide by two large posters with the message “Instead of pills, just use ear plugs” and “Instead of pills, just use a mask” (Figure 1), displayed on all wards. For patients, the Sleep-friendly Hospital Initiative provided online information about sleep hygiene and tips for sleeping better in the hospital. For hospital doctors and nurses, the hospital administration published a hospital-wide, interdisciplinary policy statement via the hospital computer system and distributed pocket-sized versions to each employee. This interdisciplinary action strategy underscored several main messages of the initiative, including the use of non-drug alternatives as a first line treatment and a medication list of appropriate medicines for older patients (Figure 2). These strategies were also highlighted in staff training courses for doctors, nurses and nursing students.

**Figure 1.**
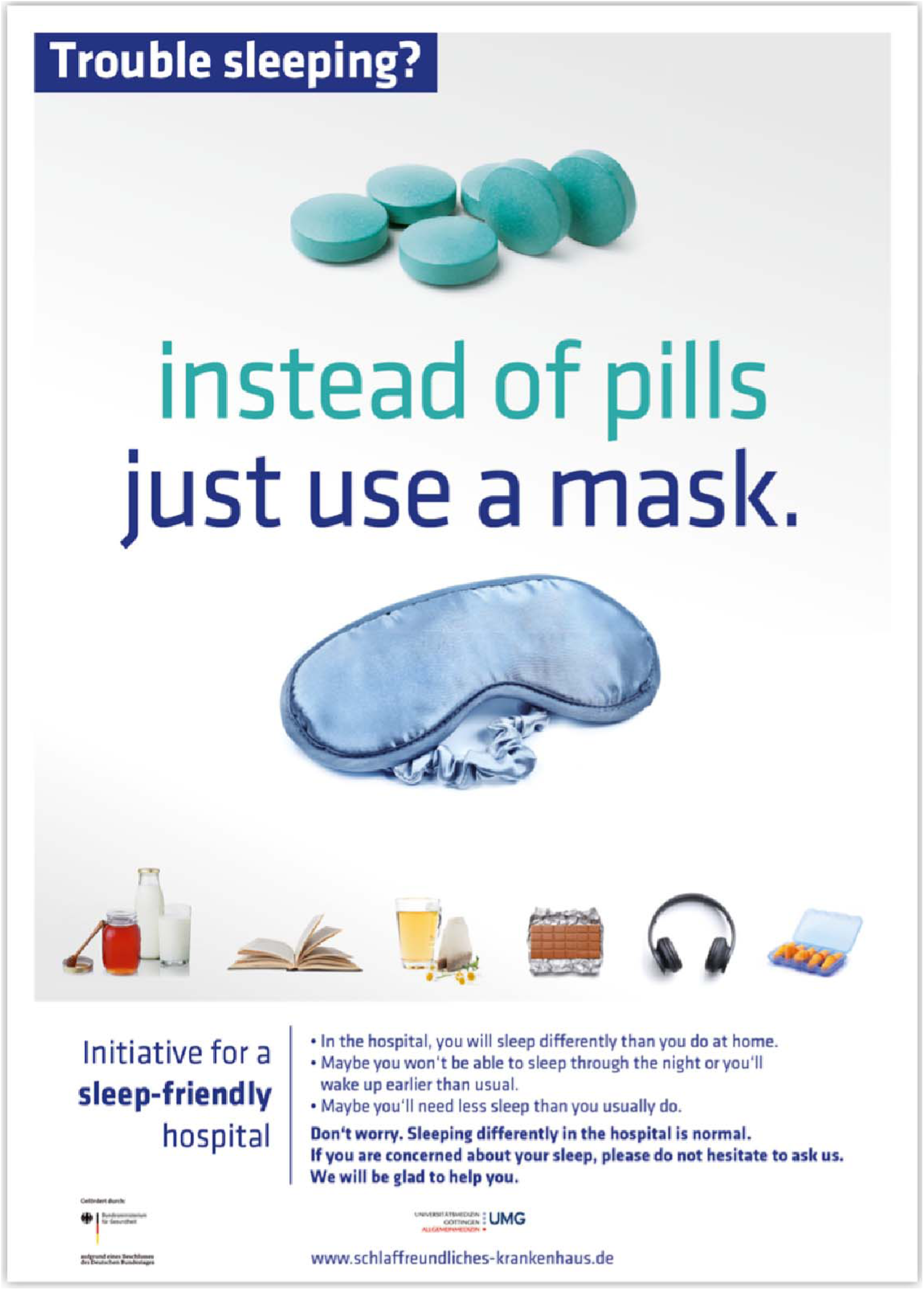
Example of a poster, highlighting alternatives to sleep-inducing drugs, which hung on every hospital ward (translation SH).

**Figure 2.**
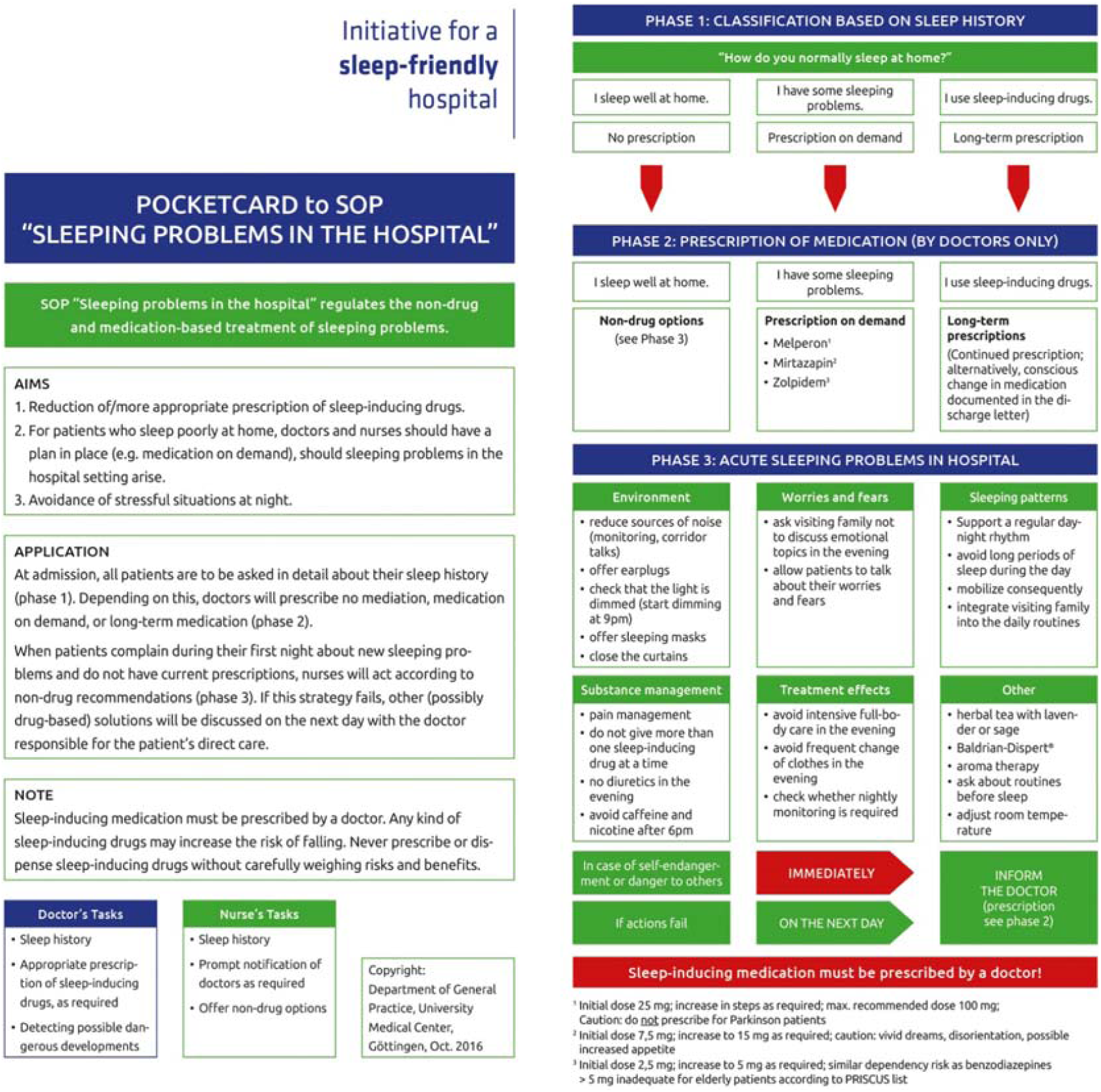
Front and back side of the hospital SOP in pocket card format (translation SH).

All stages of the implementation of the intervention were supported by the relevant stakeholders, such as hospital directors, administrators, medical and nursing staff and employee representatives.

### Comparison of prescription data before and after the intervention

The data from 960 patients aged 65 and older before intervention and 1049 patients after intervention were analysed. The percentage of female patients in 2013 and 2017 was nearly identical (58.1% vs. 57.8%; Table 1), also the mean age of the patients (79.4 vs. 79.9). In both periods, most patients (41.2% vs. 40.8%) were treated on surgical wards. Somewhat more patients had at least one psychiatric or neurological disorder in the pre-intervention period (24.1% vs. 22.3%). Slightly fewer patients entered the hospital without pre-existing sleep-inducing drug use in the pre-intervention period (67.1% vs. 68.3%).

**Table 1.** Characteristics of the hospitals’ patients.

| **Baseline characteristics** | **2013**<br>(960 patients) | **2017**<br>(1049 patients) |
| --- | --- | --- |
| **Sex** |  |  |
| Female (%) | 58.1 | 57.8 |
| Male (%) | 41.9 | 42.2 |
| **Age group** |  |  |
| 65 to 84 years (%) | 71.8 | 70.7 |
| 85 years and older (%) | 28.2 | 29.3 |
| **Type of ward** |  |  |
| Internal medicine (%) | 39.9 | 37.1 |
| Surgical departments (%) | 41.2 | 40.8 |
| Geriatrics (%) | 19.0 | 22.1 |
| **Psychiatric/neurological disorder** |  |  |
| Any diagnosis (%) | 24.1 | 22.3 |
| Dementia (%) | 13.9 | 12.3 |
| Depression (%) | 9.7 | 8.2 |
| Anxiety or panic disorder (%) | 1.4 | 2.1 |
| Sleeping disturbance (%) | 0.6 | 0.6 |
| **Sleep-inducing medication at admission** |  |  |
| None (%) | 67.1 | 68.3 |
| Regular/daily use (%) | 21.8 | 19.8 |
| Use as needed (p.r.n prescription) (%) | 3.8 | 4.5 |
| Regular and additional "as needed" (%) | 3.5 | 3.2 |
| No documentation (%) | 3.9 | 4.2 |
| **Where the patient came from** |  |  |
| Home (%) | 66.3 | 67.9 |
| Nursing home (%) | 14.7 | 12.6 |
| Other hospital (%) | 6.2 | 5.3 |
| Other department of same hospital (%) | 12.6 | 12.5 |
| Unknown (%) | 0.3 | 1.7 |

### Use of sleep-inducing drugs

Before intervention, 483 (50.3%) of the patients received sleep-inducing drugs at some time during their hospital stay. After the intervention, 381 (36.3%) patients received a sleep-inducing drug, a significant reduction by 14 percentage points (OR = 0.56; 95% CI: 0.47 to 0.68; p < 0.001; Table 2). The reduction was particularly pronounced in patients without a psychiatric or neurological disorder: from 45% (328/729) to 28.8% (235/815), a reduction of 16.2 percentage points, compared to a reduction of 4.7 percentage points in those patients with a psychiatric or neurological disorder (from 67.1% to 62.4%).

**Table 2.**
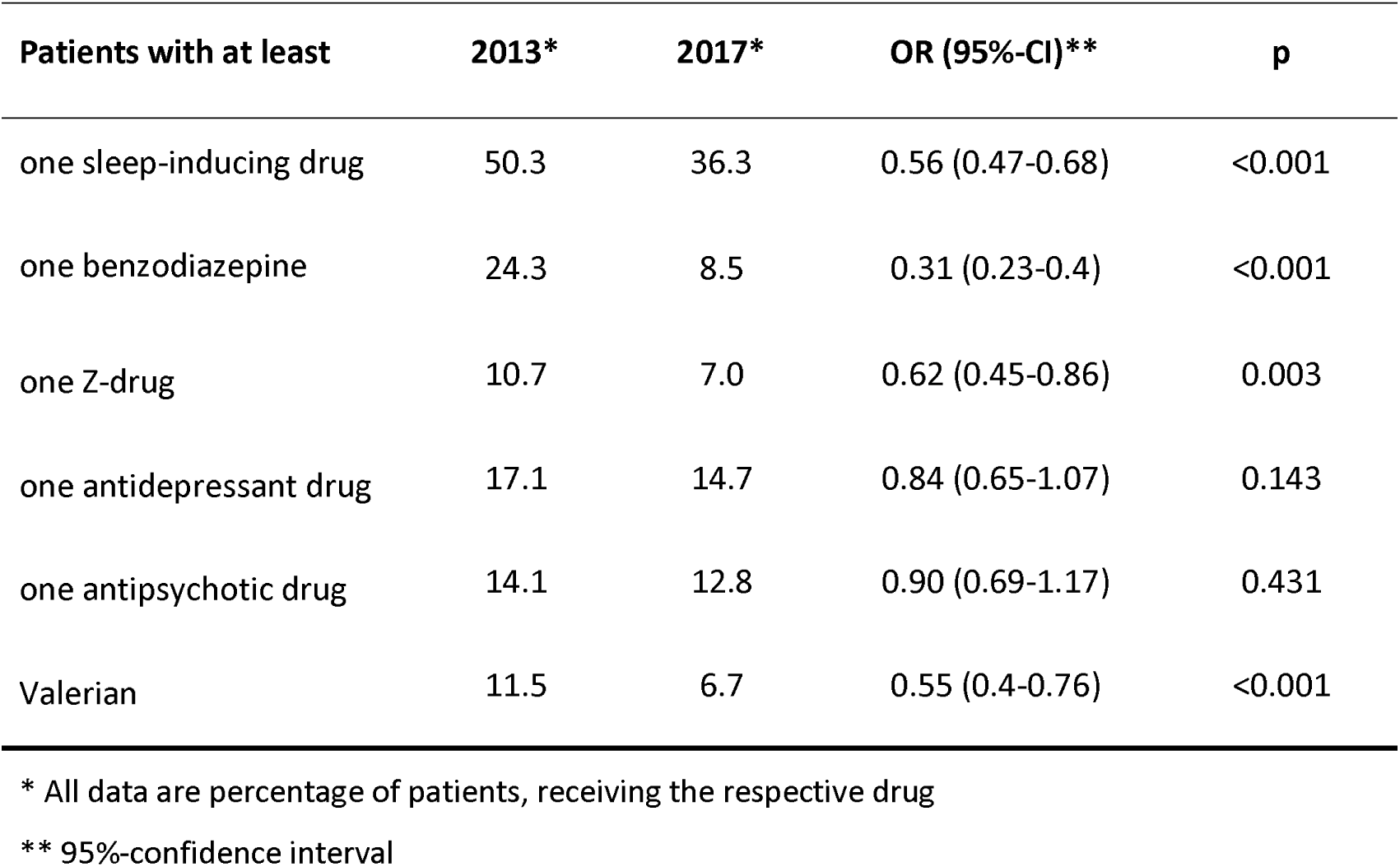
Change between pre- and post-intervention period

### Benzodiazepines

Prior to the intervention, one quarter (24.3%) of all older hospital patients received benzodiazepines (Table 2). After the intervention, this percentage was reduced by 15.8 percentage points to 8.5% (p < 0.001).

### Z-drugs

The percentage of patients treated with Z-drugs dropped between the first and second measurement (10.7% to 7.0%; 3.7 percentage points; p = 0.003).

### Other sleep-inducing drugs

The percentage of patients treated with sedative anti-depressants was slightly reduced from 17.1% before the intervention to 14.7% after the intervention (2.4 percentage points). Anti-psychotic treatment, too, declined slightly (from 14.1% to 12.8%). Sleep-inducing drugs were not substituted by valerian on a large scale; on the contrary, valerian itself was reduced from 11.5% to 6.7% (p < 0.001).

## DISCUSSION

### Summary of main findings

We observed a significant decline of sleep-inducing drugs in the intervention hospital. The prescription of benzodiazepines and Z-drugs was reduced without increasing the use of alternatives such as sedative anti-depressants, anti-psychotics or valerian.

### Strengths and limitations of the study

A strength of the study is that we introduced the intervention in, and analysed its effect for, the entire hospital and not only on selected wards as in many other studies [17, 18]. In these studies, the participating doctors and nurses may have gone the extra mile not mainly because of the intervention but because they felt as pioneers and being observed and singled out (Hawthorne effect) [19].

Further sub-analyses helped figure out whether the intervention reached the target drugs (benzodiazepines and Z-drugs) and the target population (patients without specific psychiatric/neurological diagnoses) without encouraging substitution with other sedative drugs.

The most important limitation of the study is the implementation of the intervention in only one hospital. Results of a single-hospital study can only be a starting point for future research and are generalizable for other settings only to a limited degree. Moreover, the study was embedded in a quasi-experimental design[20] where the intervention was not randomly assigned. So, in principle, we cannot exclude confounding.

### Comparison with the literature

#### Overall reduction of sleep-inducing drugs

We witness an increasing sensitivity especially towards the use of benzodiazepines during the last decades, as can be seen by the German guidance for medical practice entitled “Medical drugs—harmful use and dependency” published in 2007 [21], also reported from 2010 onwards in annual prescription statistics from office-based physicians, for example, in the German Arzneiverordnungs-Reports [22]. However, the decline in the intervention hospital clearly outperformed the decline reported in these prescription statistics by a factor of around 2.

Several research projects, mainly in English speaking countries, have tried to reduce sedative-hypnotic use in hospitals[23], with reduction rates between 3 and 19 percentage points. For example, a study in a UK hospital which improved the sleep environment, especially the noise level, could reduce the percentage of as-needed sedatives from 37% to 16% [18]. A computer-based reminder in a Connecticut academic healthcare centre reduced the prescribing of sedative and hypnotic drugs from 18% to 12% of hospitalized older patients [24]. Our intervention is, to the best of our knowledge, the first one in German and its success is at the upper end of similar studies.

#### Targeted reduction of benzodiazepines and Z-drugs

The relevant patient safety concerns when prescribing benzodiazepines and Z-drugs for older patients include confusion, falls, fractures and craving[4]. Previous work showed that benzodiazepines were being prescribed often—and in too high doses—to older patients in the intervention hospital [25]. Therefore, the intervention components of the Sleep-friendly Hospital Initiative were designed to change the habit of prescribing benzodiazepines to older patients with sleeping problems and offer safer alternatives in line with the PRISCUS list, such as non-drug alternatives, valerian, melperone, mirtazapine and low-dose zolpidem. As intended, the percentage of older patients receiving benzodiazepines was significantly reduced in the intervention hospital. In addition, the listing of zolpidem on the recommended list of drugs in the hospital-wide SOP did not increase the use of Z-drugs in the intervention hospital. Rather, the prescription rate of Z-drugs modestly declined over time to a similar degree. In contrast to many other studies [18, 26], our analysis controlled whether psychotropic drugs were not strongly reduced in patients who may benefit from them. We could show that the reduction, indeed, addressed the target persons, i.e., those without a psychiatric or neurological disease.

#### The role of hospital pharmacists

Although hospital pharmacists are important in the clinical support of patients and pharmacist-delivered interventions in inpatient settings can lead to significant improvements, as most recent studies have shown [27, 28], they were not in the focus of our study. One reason is that in Germany, especially in the intervention hospital under study, hospital pharmacists often do not play a leading role when it comes to changes in pharmacological management. However, the results of our study could be of interest to hospital pharmacists who are currently searching for a cost-effective strategy to improve drug safety for older hospitalized patients [29].

## CONCLUSIONS

A multi-faceted intervention to change the practice of the use of sleep-inducing drugs in a German hospital proved to be successful in terms of drug reduction. Before we recommend similar interventions and strategies for other hospitals, we should consider some prerequisites for being successful.

Since sleep problems are seldom at the top of the list of concerns in an average hospital, clear leadership by hospital directors is needed. These leaders must be committed and willing to champion this project by repeatedly emphasizing the importance of first-line non-drug treatment. Our intervention may provide inspiration to those searching for a strategy to improve drug safety for older hospitalized patients. However, every institution must find its own way, i.e. adapt the intervention to the hospital and its staff. Such participatory development of an intervention seems to be vital for the success of the intervention. The results of this pilot study give an impression of the interventions’ possible impact and provide essential information for the design of a randomized controlled study.

## Data Availability

On reasoanable request

## Acknowledgements

Our special thanks goes to Michael Karaus (Evangelisches Krankenhaus Göttingen-Weende), the director of the participating hospital, who gave access to patient health records documenting the prescription and use of sleep-inducing drugs, diagnoses, etc. The intervention would not have been possible without the whole-hearted participation of the doctors, nurses and administrators of the intervention hospital. The results presented here represent the culmination of a five-year-long research project, which was only possible with the hard work of several researchers, PhD students and Masters-level students: Inken Arnold, Lea Kauffmann, Freya Neukirchen, Katharina Schmalstieg-Bahr, Kati Straube, Sven Wedeken, Fabian Wedmann and Vivien Weiß. We thank our study nurses, Simone Assmann, Anouschka Kausche, Sara Krömer and Ida Wilkens, for their diligent work documenting the hospital records needed for this data analysis. We also thank Daniela Keller, whom we consulted about the power calculation and statistical analysis.

## Ethics approval

The study was approved by the Ethics Committee of the University Medical Center in Göttingen, Germany (ref number 25/2/14).

## Contributorship

All authors designed the study; JK collected the data; SH, JK and WH performed all statistical analyses; SH and WH were the major contributors in writing the manuscript and are the guarantors of the paper. All authors read, discussed and approved the final manuscript.

