## Supplement Table 1 for "Reducing the use of sleep-inducing drugs during hospitalization by a multi-faceted intervention – a pilot study"

Appendix. List of sleep-inducing drugs

**Benzodiazepines**

Lormetazepam

Lorazepam

Oxazepam

Diazepam

Temazepam

Other Benzodiazepines

**Z-Drugs**

Zopiclone

Zolpidem

**Anti-psychotics**

Melperone

Quetiapine

Prothipendyl

Promethazine

Haloperidol

Risperidone

Other Anti-psychotics

**Anti-depressants**

Mirtazapine

Amitriptyline

Doxepin

Other Anti-depressants

**Other sleep-inducing drugs**

Valerian
